# Oat protein nanofibril-iron hybrids as a stable, high-absorption iron delivery platform for human nutrition

**DOI:** 10.1101/2025.01.24.25321072

**Authors:** Jiangtao Zhou, Sueppong Gowachirapant, Christophe Zeder, Alexander Wieczorek, Ines Kutzli, Sebastian Siol, Ferdinand von Meyenn, Michael B. Zimmermann, Raffaele Mezzenga

**Affiliations:** Laboratory of Food and Soft Materials, Department of Health Sciences and Technology, ETH Zurich, Zurich, Switzerland; Institute of Nutrition, Mahidol University, Nakhon Pathom, Thailand; Laboratory of Human Nutrition, Department of Health Science and Technology, ETH Zurich, Zurich, Switzerland; Laboratory of Nutrition and Metabolic Epigenetics, Department of Health Science and Technology, ETH Zurich, Zurich, Switzerland; Laboratory for Surface Science and Coating Technologies, Empa – Swiss Federal Laboratories for Materials Science and Technology, Duebendorf, Switzerland; Department of Medical and Molecular Genetics, King’s College London, London, UK; Translational Immunology Discovery Unit, MRC Weatherall Institute of Molecular Medicine, John Radcliffe Hospital, University of Oxford, Oxford, UK; Department of Materials, ETH Zurich, Zurich, Switzerland

## Abstract

Iron deficiency and anemia are significant global health issues, affecting nearly two billion people worldwide. The World Health Organization recommends iron fortification of foods as an effective strategy to reduce anemia rates. Available iron fortificants, however, are limited by low absorption and/or poor sensory performance. Developing new iron compounds that deliver highly bioavailable ferrous iron in foods without compromising sensory quality remains a challenge. In this work, we introduce oat protein nanofibrils (OatNF) carrying ultrasmall iron nanoparticles as promising iron fortificants for foods and beverages. Tailored synthesis of OatNF hybrids can produce iron nanoparticles in either the ferrous or ferric state. When sodium ascorbate (SA) is used as the reducing agent, the OatNF carry stabilized ferrous iron which is remarkably well absorbed in humans. In iron-deficient women, geometric mean absorption is found to be 46.2% (95% CI: 39.1%–54.7%) when given with water and 13.4% (95% CI: 9.8%–18.3%) when given with a polyphenol-rich food, representing 76% and 66% higher absorption, respectively, than the reference compound, FeSO_4_. When NaOH is used as the reducing agent, the OatNF carry mainly ferric iron, which is well-absorbed and shows superior sensory performance in reactive food matrices. These promising results introduce OatNF hybrids as a possible cost-effective, plant-based and organoleptic-friendly solution to the global challenge of iron deficiency anemia.

## Introduction

The worldwide prevalence of anaemia across all age groups is 24.3% (95% UI 23.9– 24.7), affecting approximately 1.92 billion people^1–4^. Dietary iron deficiency is the leading cause of anaemia-related years lived with disability (YLDs), with a YLD rate of 422.4 (95% UI 286.1–612.9) per 100,000 population^1^. In order to reduce anemia, the World Health Organization (WHO) recommends use of ferrous iron salts for fortification of most foods^5^, but most iron fortificants remain poorly effective due to low bioavailability and poor sensory performance^2,3,6–9^, so that developing a robust technology capable to deliver highly bioavailable iron in foods without causing negative sensory effects is still highly sought after^8,10,11^. Consequently, although the WHO has set a global nutrition target to reduce anaemia prevalence by 50% among women of reproductive age (15–49 years) by 2030^12^, the global prevalence of anaemia in this group of women remains stubbornly high, at 30% (27–33%)^13^. Symptoms of anaemia in women include fatigue, weakness, difficulty concentrating, dizziness, pallor, and headache^8^.

The recommended daily iron intake for young women is 18 mg^14^, but this high requirement is most often not met through diet alone. To address this, iron fortification of staple foods is the recommended strategy to increase iron intake among young women^5,6^. WHO recommends use of ferrous iron salts for fortification of most foods, with ferrous sulphate (FeSO_4_) considered the gold standard due to its high bioavailability and low cost^5^. However, incorporating ferrous iron into many foods is challenging because it typically causes undesirable sensory changes^5^. Furthermore, iron added to food is often poorly absorbed because of inhibitory compounds, particularly polyphenols and phytic acid^6^. Finally, low iron absorption from fortificants also results in large amounts of unabsorbed luminal iron that may provoke gut inflammation and dysbiosis^15,16^. To partially address this, compounds like sodium iron EDTA (NaFeEDTA) have been developed, offering better absorption compared to ferrous salts when used in food matrices with inhibitory compounds^5^. Yet, despite these advances, finding a method to deliver highly bioavailable iron in foods without causing negative sensory effects is still highly sought after, and remains both a significant challenge and a key area of research.

Here, we introduce oat protein nanofibrils (OatNF) carrying ultrasmall iron nanoparticles as a promising novel strategy to provide highly bioavailable iron with enhanced sensory performance in foods and beverages. We show that when sodium ascorbate (SA) is used as a reducing agent of iron salt precursors in the presence of OatNF, sub-nm nanoparticles of ferrous iron form on the surface of the OatNF (OatNF-SA-Fe), producing hybrids with superior iron delivery features. OatNF exhibited strong iron-binding, reducing and stabilizing properties during the synthesis of OatNF-SA-Fe hybrids, resulting in exceptional ferrous iron stability.

In a prospective crossover study in iron-deficient Thai women (n=52), we used erythrocyte incorporation of oral stable iron isotopes^17^ to quantify the iron absorption and bioavailability from OatNF-SA-Fe hybrids. Absorption from the hybrids was quantified when given with water alone and with a highly inhibitory polyphenol-rich meal, and compared to FeSO_4_.

The OatNF-SA-Fe hybrids were highly absorbed, with a geometric mean absorption rate of 46.2% (95% CI: 38.9%–55.0%) when administered with water, 76% higher than absorption from FeSO4, which had an absorption rate of 26.3% (95% CI: 21.4%– 32.4%). Similarly, when administered with a polyphenol-rich meal, the iron absorption from the OatNF-SA-Fe hybrids was 67% greater than from FeSO_4_. We infer that the combination of OatNF and sodium ascorbate (at a 2:1 molar ratio to iron) enhances iron absorption, likely due to their reducing and stabilizing effects, as well as the relatively high content of glutamine in the OatNF. This conclusion is further supported by experiments using NaOH-chelated hybrids (OatNF-NaOH-Fe), which, in contrast to the OatNF-SA-Fe hybrids, contain high ferric iron content. These hybrids were also well-absorbed and achieved 80% and 75% bioavailability compared to FeSO_4_ when administered with water and a polyphenol-rich meal, respectively. Additionally, the OatNF-Fe hybrids demonstrated excellent organoleptic qualities, showing minimal sensory impact when added to common foods and beverages.

These findings build on our previous research, in which we first proposed the use of nanosized iron as an iron fortification strategy^18^ and showed that nanosized ferric phosphate is well absorbed in both mice and humans^19^. Yet, in those studies, iron bioavailability remained lower than that of FeSO_4,_ and, in humans, only achieved 72% relative bioavailability to FeSO_4_. We also demonstrated the high bioavailability of iron nanoparticle-milk-derived protein nanofibrils in rats^20^ and confirmed the safety of food amyloid fibrils as nutritional ingredients through in vitro and in vivo assessments^21^. While these nanofibril studies were limited to mouse models and used animal-derived proteins, they provide the foundation to the present work, where iron bioavailability in humans achieved 176% relative bioavailability to FeSO_4_. Given the above, we believe that the plant-based OatNF-SA-Fe hybrids introduced in this work may offer a breakthrough strategy for effective delivery of highly absorbed iron in fortified foods and beverages and may contribute to the mitigation of iron deficiency and anemia on a global scale.

## Results and discussion

### The fabrication of our OatNF-SA-Fe hybrid

The preparation process of OatNF-SA-Fe hybrid is shown in Fig. 1a. Initially, oat globulin (OG), the primary oat protein, was extracted from oat flake powder and then was incubated to fabricate OG nanofibrils under acidic condition (pH 2) and heat treatment^22^. The colloidal dispersion of OG nanofibril was subsequently mixed with iron chloride and sodium ascorbate (SA), yielding a translucent dispersion of iron-nanofibril (OatNF-SA-Fe) hybrids.

**Fig. 1.**
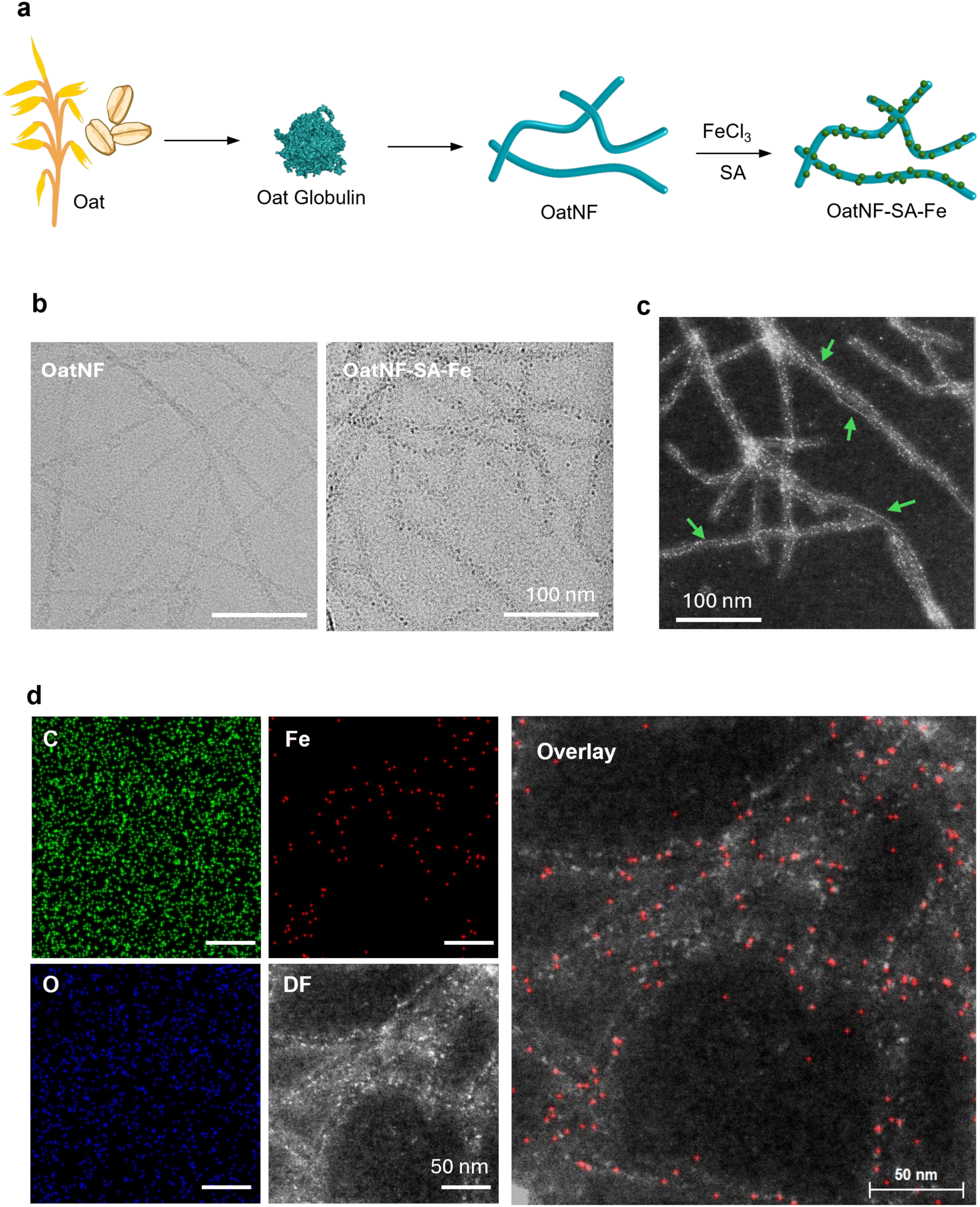
Fabrication and electron microscopy characterization of the OatNF-SA-Fe hybrid. **a**, Schematic illustration of the fabrication process of the OatNF-SA-Fe hybrid. **b**, CryoEM images of oat nanofibril and OatNF-SA-Fe hybrids before and after iron nanoparticles synthesis. **c**. HAADF-STEM image of the OatNF-SA-Fe hybrid, where the bright dots anchored and necklace-aligned (arrow) along OatNF correspond to the heavier atomic number of iron in the hybrid. **d**. EDS elemental mapping images of the OatNF-SA-Fe hybrids, showing distributions of carbon (C), oxygen (O) and iron (Fe), and the corresponding HAADF-STEM image. The overlay of HAADF-STEM image and Fe elemental map confirms the accumulation of iron along the oat nanofibrils.

Cryo-transmission electron microscopy (cryoTEM) images (Fig.1b and Supporting Fig. S1a) revealed that iron nanoparticles formed and were anchored onto the surface of OG nanofibrils. This iron-decoration was more evident using high-angle annular dark field scanning TEM (HAADF-STEM) images (Fig. 1c and Supporting Fig. S1b). Numerous bright dots along oat nanofibril are observable, representing iron particles and ascribed to the atomic number contrast between iron and amino acid (AA) elements in the protein nanofibrils. Given the observed diameter of 4-10 nm for the OatNF, these iron particles were estimated to be at most 1 nm or less in size. This nanoscale size is likely due to SA-mediated iron chelation that prevents iron aggregation into large complexes^23^. Remarkably, we also noticed that these sub-nm particles were aligned along the fibrils (Fig. 1c and Supporting Fig. S1b). The presence and distribution of iron on these nanoparticles are further validated by elemental mapping using energy dispersive X-ray spectroscopy (EDS) analysis (Fig.1d and Supporting Fig. S1c). The overlay of the Fe elemental map and the corresponding HAADF-STEM image demonstrated iron conformation in these sub-nm particles along the surface of OatNF.

To prove the role of SA-Fe chelation in forming sub-nm iron particles, we conducted a control study by mixing OatNF with FeCl_3_ and sodium hydroxide (NaOH). The CryoTEM and HAADF-TEM images (Supporting Fig. S2) showed the formation of iron nanoparticles with the sizes of tens nanometers, consistent with our previous study^24^. These particles were significantly larger than those in the OatNF-SA-Fe hybrid, indicating that NaOH-chelated iron tended to aggregate into large complexes before nucleating on the OatNF surface and forming OatNF-NaOH-Fe hybrid. Furthermore, Fourier transform infrared (FTIR) spectroscopy also confirmed SA-iron chelation during forming the iron particles in the OatNF-NaOH-Fe hybrid.

### Chemo-physical characterization of the OatNF-SA-Fe hybrid

The morphological and chemo-physical characterization of the iron complex in the OatNF-SA-Fe hybrid was carried out in detail to evaluate the conversion rate of Fe(III) into the more bioavailable Fe(II). Three complementary methodologies were employed to assess this conversion across different length-scales.

X-ray photoelectron spectroscopy (XPS) measurements were carried out to determine the chemical composition and chemical state of our OatNF-SA-Fe hybrid. Fig. 2a shows the XPS Fe 2p core level spectrum of OatNF-SA-Fe hybrid with a primary peak at 710 eV and a satellite peak at 715 eV. Remarkably, theO 1s spectrum (Fig. 2b and Supporting Fig. S3c) indicates the absence of Fe-O bonds, associated with iron oxides, iron hydroxides and oxygen ligands, which is however present for the OatNF-NaOH-Fe hybrid (Fig. 2b and Supporting Fig. S3d). Besides Na and Cl, no other contaminations were found in the survey spectrum of our OatNF-SA-Fe compound (Supporting Fig. S3a) and thus the iron in the OatNF-SA-Fe hybrid exists primarily as non-oxide Fe(II) and Fe(III) species. Fitting of the Fe 2p region (Figs. 2a and Supporting Fig. S3c) revealed a remarkably high amount (ca. (91±5)%) of the bioavailable Fe (II) in the OatNF-SA-Fe hybrid. This high conversion rate is attributed to the sub-nm iron particles anchored on OatNF surface that can be easily reduced by protein nanofibrils^20^. In contrast, the OatNF-NaOH-Fe control hybrid showed larger iron complexes on the fibril surface (Supporting Fig. S2c) and its O 1s core level spectra (Figs. 2b and Supporting Fig. S3d) indicated a clear Fe-O feature. To estimate the Fe(II) content in this complex Fe mixture, fitting of the Fe 2p was performed based on peak constraints from literature reports relating to pure Fe chlorides, oxides, and oxy-hydroxides (Supporting Fig. S3d)^25^. In agreement with our previous study^24^, the fitting result indicated approximately 30-40% of Fe(II) and remaining 60-70% iron was present as Fe (III) species.

**Fig. 2.**
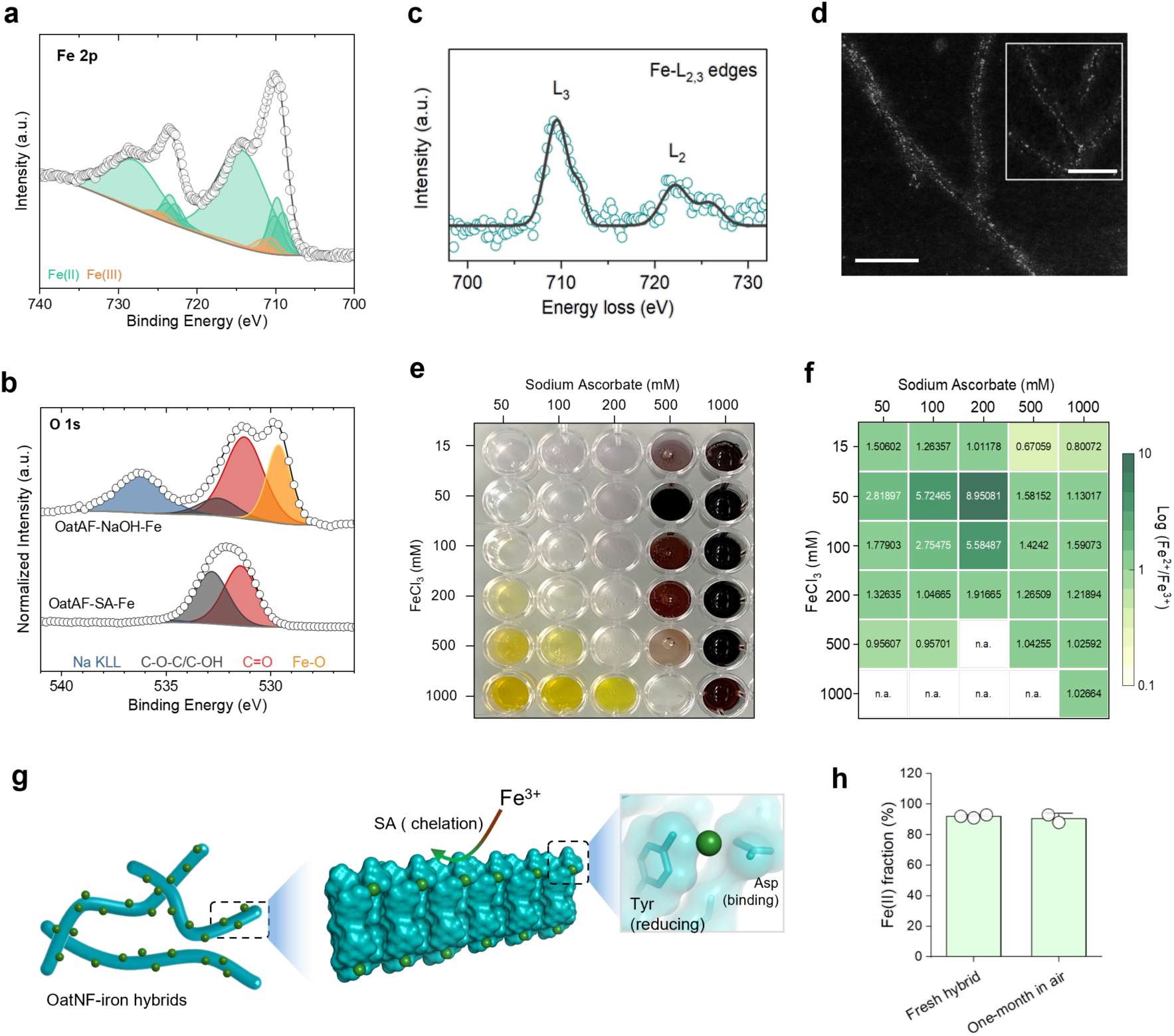
Characterization of the Oat-SA-Fe hybrids. **a-b,** XPS spectrum at representative Fe 2p region (**a**) for the OatNF-SA-Fe hybrid. Multicomponent fitting was performed according to previous literature^25^. XPS spectrum of the O 1s region (**b**) comparing the OatNF-SA-Fe and OatNF-NaOH-Fe hybrids. The Fe-O feature is clearly visible in OatNF-NaOH-Fe hybrid but is absent in the OatNF-SA-Fe hybrid. **c,** Background-subtracted EELS spectrum of the OatNF-SA-Fe hybrid at the Fe L_2,3_-edges. **d,** HAADF-TEM image of aligned iron particles on the nanofibril surface in the OatNF-SA-Fe hybrid. The scale bars are 50 nm. **e-f,** OatNF-SA-Fe dispersion at different concentrations of FeCl_3_ and SA (**e**) and the determination of Fe^2+^/Fe^3+^ ratio through the colorimetric assay by potassium ferricyanide and potassium ferrocyanide reagent (**f**). **g,** Illustration of chelation, binding, reduction, stabilization and preservation of iron particles (green) on the OatNF surface. The X-ray crystallographic structure of the amyloid-forming fragment (Ala-Val-Try-Val-Phe-Asp) of oat protein reveals that surface AAs, including aspartic acid and tyrosine, in OatNF may serve as specific sites for iron binding and reduction, respectively. **h,** The fraction of ferrous iron in freshly prepared OatNF-SA-Fe hybrid and after exposure to air for one month, demonstrating the stability of ferrous state.

Electron energy-loss spectroscopy (EELS) coupled with STEM was employed for microscale analysis of Fe chemical state in the OatNF-SA-Fe hybrid. EELS spectrum (Supporting Fig. S4o) at Fe *L*_2,3_-edges (Fig. 2c) displayed two peaks corresponding to *L*_3_ and *L*_2_ edges with a line separation of 13.2 eV. The Fe L_3_ edge exhibited two sub-peaks, that are a main peak (709 eV) with a minor shoulder at 711 eV referring to ferrous and ferric content respectively^26,27^. The EELS *L*_3_/*L*_2_ intensity ratio^28^ is related to Fe valence state (*Fe^3+^/∑Fe*) and thus Fe^3+^ to Fe^2+^ conversion^26,29^, revealing that only 11.7% of the total iron was in the ferric state (Fe³⁺). The HAADF-STEM images (Fig. 2d) demonstrated that rich iron complexes were well-aligned along OatNF surface, confirming the specific OatNF-iron chelation that likely facilitate the reduction of sub-nanometer iron complexes to a high ferrous content.

The valence state of iron in the Oat-SA-Fe dispersion was investigated using a bulk colorimetric assay. Dispersions were first prepared at varying molar ratios of FeCl_3_ and SA, as shown in Fig. 2e. In the case of excess FeCl_3_, the yellow dispersion indicated a ferric-dominant composition. Conversely, excess SA at the high SA-to-Fe ratio tend to chelate iron to form large insoluble complexes^30^ in the mahogany-colored suspension. Remarkably, when the SA:FeCl_3_ ratio approached 2:1, the dispersion became translucent, indicating the absence of large aggregates nor concentrated FeCl_3_. To quantify the iron valence states, we measured ferric and ferrous concentration via the bulk colorimetric assay by potassium ferricyanide and potassium ferrocyanide reagents respectively (Supporting Fig. S4 a-d) and thus calculated Fe^2+^/Fe^3+^ ratio (Fig. 2f). This assay confirmed that extreme SA-to-Fe ratios, either high or low, resulted in low Fe^2+^/Fe^3+^ ratios, likely due to SA-Fe chelation or SA depletion. Remarkably, an appropriate and optimized SA-to-Fe ratio at low concentrations achieved a high ferric-to-ferrous conversion. Specifically, a high Fe^2+^/Fe^3+^ ratio of 5.6:1 (85% Fe^2+^) was observed in our synthesized Oat-SA-Fe hybrid with 100 mM FeCl_3_ and 200 mM SA, enabling its superior performance. We further investigated the ferric and ferrous content in our Oat-SA-Fe dispersion via the colorimetric assay by using ferrozine and ammonium thiocyanate respectively (Supporting Fig. S4 e-n). The assay confirms the high proportion of ferrous content and the lack of ferric content in our synthesized Oat-SA-Fe solution.

The synthesis process of the OatNF-SA-Fe hybrid is illustrated in Fig. 2g. An appropriate concentration of SA facilitates mild SA-iron chelation^23^ and then sub-nm iron particles before binding onto OatNF surface. The alignment of sub-nm iron particles (Figs. 1c-d, 2d and Supporting Fig. S1b) indicated a specific iron-binding site of OatNF. The major binding sites are mostly likely the carboxyl groups of glutamic and aspartic acids^31,32^, which are abundant in oat globulin (Supporting Fig. S4p), to form highly stable iron complexes with tridentate structure^32,33^. Subsequently these immobilized iron nanoparticles were reduced by the reducing AAs present on the surface of nanofibrils, including cysteine, tryptophan and tyrosine^20^. Among them, tyrosine is particularly abundant in oat protein. Fig. 2g also highlights the molecular structure of amyloid-forming fragments (Ala-Val-Try-Val-Phe-Asp) in oat protein, illustrating the sites on oat nanofibril where aspartic acid facilitates iron-binding and tyrosine may contribute to the reduction of iron nanoparticles. This chelating-binding-reducing process achieves a high (90%) ferric-to-ferrous conversion. Furthermore, OatNF exhibits strong antioxidant property to stabilize and preserve iron complexes in the bioavailable ferrous state. The freeze-dried OatNF-SA-Fe hybrid demonstrated exceptional long-term stability, retaining a high ferrous iron content even after being exposed to air for one month (Fig. 2h and Supporting Fig. S3e). This stability makes the OatNF-SA-Fe hybrid suitable for practical applications in the iron fortification of foods.

### Sensory study in fortified foods and beverages

We next assessed the sensory performance of the OatNF-SA-Fe and Oat-NaOH-Fe hybrids when added to foods and beverages, compared to two commonly used iron fortificants: FeSO_4_, the reference compound, and ferric pyrophosphate, a sensorially-inert but poorly absorbed compound. To quantify their sensory impact, we measured the color changes induced by the four iron compounds two hours after their addition to bottled drinking water, apple juice, chocolate-flavored whole milk, strawberry yogurt, and wheat- and maize-based porridges. All compounds were added to the food and beverages at a fortification level of 2 mg iron per serving.

Fig. 3 and Supporting Fig. S5 show the sensory performance of the compounds was matrix dependent. The relatively inert ferric pyrophosphate generally caused the least color change. In apple juice and wheat porridge, the addition of both the Oat-NaOH-Fe and Oat-NF-SA-Fe resulted in minimal color changes that were below the limit of detection. In general, FeSO_4_ caused the greatest changes in color, followed by the Oat-NF-SA-Fe. These results are likely explained by the differing solubility of iron in the different compounds. While Oat-NF-SA and ferrous sulfate are fully soluble and can therefore easily react with food components like polyphenols, producing colored complexes, ferric pyrophosphate is very poorly water soluble, and its iron is not available to react. Oat-NaOH-Fe contains both soluble iron and insoluble particles, resulting in color changes between the soluble and insoluble compounds. The color change in water for all compounds but ferric pyrophosphate is likely due to the oxidation of Fe(II) to Fe(III) at neutral pH in the presence of oxygen, resulting in the formation of suspended reddish ferric hydroxide. Importantly, we note that, when dissolved in water and tasted by the investigators, the OatNF-SA-Fe solutions exhibited a pleasant oat flavor and aroma, enabling administration in water without the need for flavour masking.

**Fig. 3.**
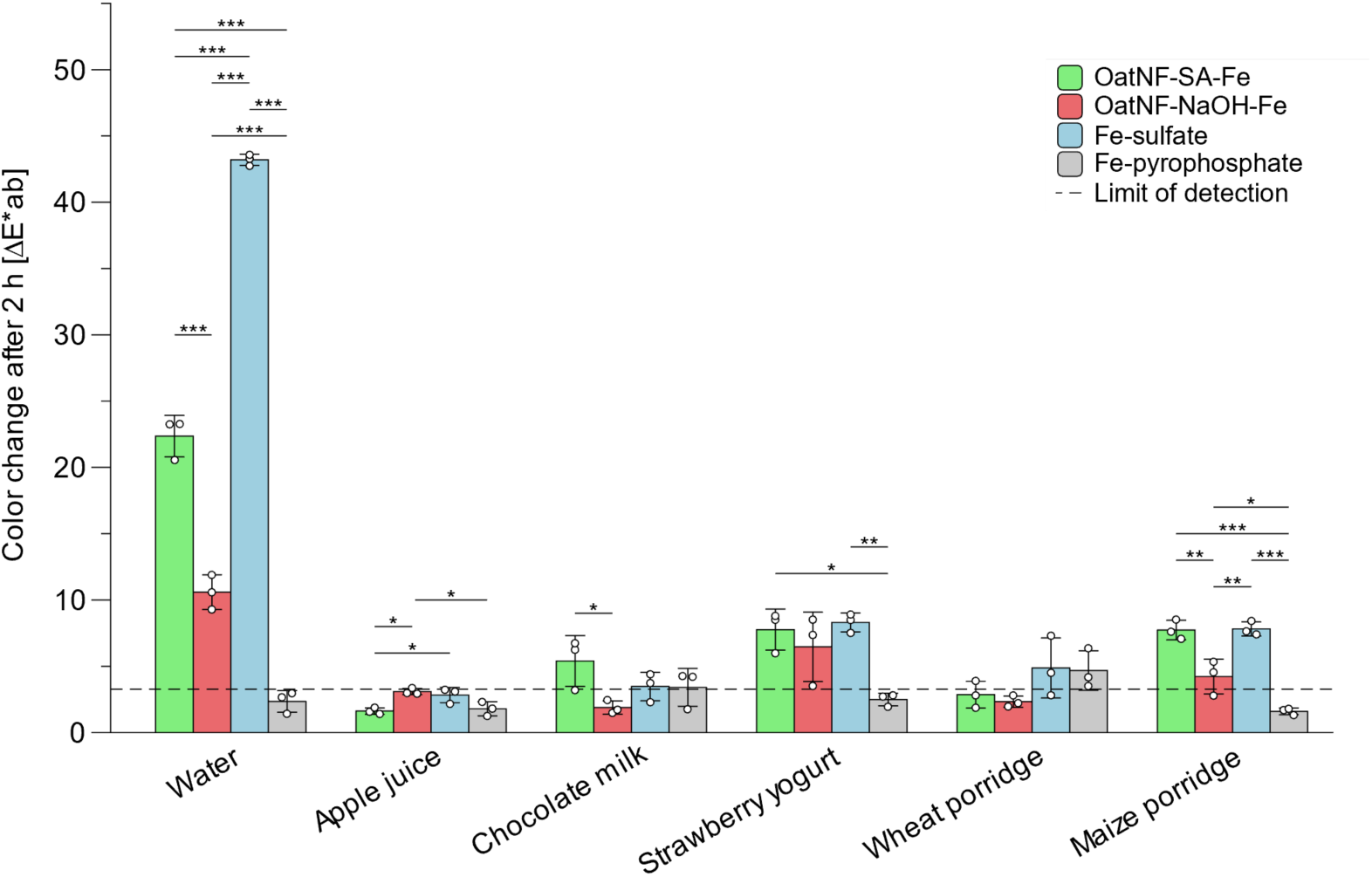
Sensory performance of Oat-SA-Fe, Oat-NaOH-Fe, ferrous sulfate and ferric pyrophosphate in various food matrices, 2 hours after mixing. The sensory performance of the OatNF-SA-Fe and Oat-NaOH-Fe hybrids compared to two commonly used iron fortificants: FeSO_4_ and ferric pyrophosphate, after their addition to bottled drinking water, apple juice, chocolate-flavored whole milk, strawberry yogurt, and wheat- and maize-based porridges, two hours after the mixing. N=3 replicates. The data are shown as mean values ± standard deviation. \**P* < 0.05, \*\**P* < 0.01, \*\*\**P* < 0.001.

### Clinical iron absorption study in iron-deficient Thai women

We conducted a prospective, cross-over study in young iron-deficient women in Nakhon Pathom, Thailand, a region in Southeast Asia where women are at high risk of iron deficiency anemia^1^. To measure iron absorption from the different iron compounds, we used a multiple iron stable isotope technique, allowing each participant to act as her own control. The method is based on the incorporation of isotopic labels into erythrocytes two weeks after oral administration^17^.

As shown in Fig. 4a, the 37-day study was designed so that each participant received six different dietary conditions. The three iron compounds tested were: (i) ^57^Fe-labelled Oat-NF-SA-Fe hybrid, (ii) ^58^Fe-labelled Oat-NaOH-Fe hybrid, and (iii) ^54^Fe-labelled ferrous sulfate (FeSO4) as the reference compound. Each dose contained 4 mg of elemental iron and all compounds were intrinsically labeled with stable isotopes during synthesis. The compounds were administered either in water or, to provide a polyphenol-rich inhibitory food matrix, mixed into açai puree with honey (Fig. 4b,c). The primary outcome was fractional iron absorption, and the pre-specified comparisons were both hybrids versus FeSO_4_ in each of the two matrices.

**Fig. 4.**
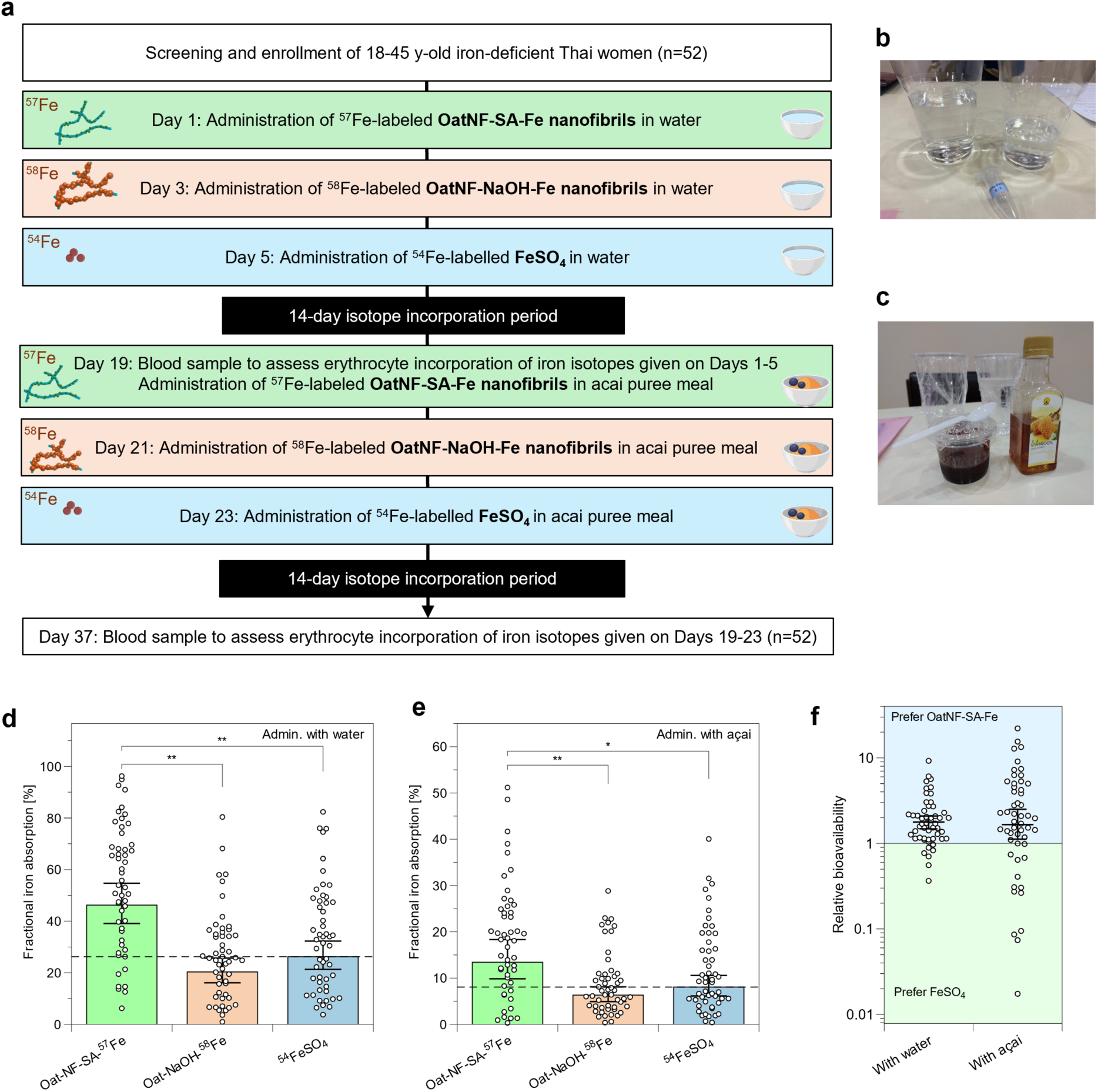
Clinical study to evaluate iron absorption from OatNF-SA-^57^Fe, OatNF-NaOH-^58^Fe and ^54^FeSO_4_ in iron-deficient Thai women. **a**, Outline of the randomized-order, cross-over study design. **b-c**, OatNF-SA-Fe compounds dissolved in water or mixed in açai puree before administration **d-e**, Fractional iron absorption in water from OatNF-SA-Fe, OatNF-NaOH-Fe and FeSO_4_ fortified foods, showing 46.2% (39.1-54.7%), 20.3% (16.1-25.7%) and 26.2% (21.3-32.3%) respectively (**d**), and 13.4% (9.8-18.3%), 6.3% (4.9-8.1%) and 8.1% (6.1-10.6%) respectively when administered with açai puree (**e**). Data are presented as geometric means with 95% confidence intervals (n=52). Compared using two-sided paired t-tests with Bonferroni adjustment for multiple testing. Dashed lines indicate the values of FeSO_4_ reference. Two absorption values >60% in the first condition are not shown to improve figure clarity. (**f**) Relative bioavailability of OatNF-SA-Fe compared to FeSO_4_ was 1.78 (1.48-2.1) when administered with water and 1.66 (1.12-2.51) with açai puree, given as geometric means (95% confidence interval). \**P* < 0.05, \*\**P* < 0.01.

During the screening phase, women aged 18–45 years with serum ferritin (SF) concentrations < 50 µg/L (n=52) were recruited. Detailed inclusion and exclusion criteria are provided in the Methods section. The baseline characteristics of the subjects are shown in Supporting Table S1. in Eligible participants received the three iron compounds with water on study days 1, 3, and 5. After a 14-day period to allow the iron isotopes to incorporate into erythrocytes, we collected venous blood samples to assess iron absorption from the first three conditions. Participants then received a second round of conditions, with the three iron compounds administered mixed into polyphenol-rich açai puree with honey on study days 19, 21, and 23. The order of the conditions was randomized for each participant for each block using a Python script. The sequences were generated so that each isotope was used only once in each block. Assignment was not masked. After another 14 days, we again collected venous blood samples to measure iron absorption from the final three conditions. In total, 312 measures of iron absorption were conducted across the 52 participants.

Fig. 4d shows iron absorption from the three compounds in water. Geometric mean fractional iron absorption (FIA) from FeSO_4_ was 26.3% (95% CI: 21.4%–32.4%). The OatNF-SA-Fe hybrid was significantly better absorbed than FeSO_4_, exhibiting a high FIA of 46.2% (95% CI: 38.9%–55.0%). Notably, iron from the OatNF-SA-Fe hybrid was 76% better absorbed (*i.e.* 1.76 times) than iron from FeSO_4_, which is fully water soluble and considered the reference compound for iron absorption. This finding may be due to the very large specific surface area (SSA) of the sub-nm sized ferrous iron in the OatNF-SA-Fe hybrid, which allows for rapid dissolution at gastric and duodenal pH during digestion, allowing greater uptake by the divalent metal transporter (DMT)-1 ^18^. In addition, the sodium ascorbate (at a 2:1 molar ratio to iron) likely enhances iron absorption, due to its reducing effect^34^. The OatNF-NaOH-Fe hybrid showed a geometric mean FIA of 20.3% (16.1-25.7%). This corresponds to 78% relative bioavailability compared to FeSO_4_ (*i.e.* 0.78 times) which is considered a moderate-to-high iron bioavailability value for potential iron fortificants^5^. In contrast to the OatNF-SA-Fe hybrid, up to 60% of the iron in the OatNF-NaOH-Fe hybrid is in the ferric form (Supporting Fig. S3d), and this includes very poorly water-soluble iron hydroxide and iron oxides, which typically exhibit absorption rates of <5% relative to FeSO_4_^5^. The high absorption of the sub-nm sized ferric iron in the OatNF-NaOH-Fe hybrid is also likely due to its very large SSA allowing for rapid dissolution at low gastric pH, as previously demonstrated for non-hybrid sub-nm sized iron oxides and phosphates^18^. Finally, the presence of OatNF may have contributed to the high FIA of both hybrids due to the high glutamine content in oat protein, comprising up to 13% of the amino acid content (Supporting Fig. S4p). Glutamine has previously been shown to enhance iron absorption *in vivo*^35^.

Dietary polyphenols strongly inhibit iron absorption by forming unabsorbable complexes with iron in the gut lumen^36,37^. Previous studies have reported reductions in iron absorption from foods and supplements due to polyphenols ranging from 40% to 90% ^36–40^. Our findings agree with these studies: the polyphenol-rich açai puree meal reduced iron absorption (Fig. 4e) from the three iron compounds by approximately 70% compared to the water condition. However, absorption from the OatNF-SA-Fe hybrids remained high at 13.4% (95% CI: 9.8%–18.3%), which was 66% higher than absorption from FeSO_4_, which was 8.1% (95% CI: 6.1%–10.6%). The higher absorption from the OatNF-SA-Fe hybrids may be partly attributed to the presence of sodium ascorbate in the hybrids, as ascorbic acid is known to reduce the inhibitory effects of polyphenols on iron absorption^41^. This suggests that the OatNF-SA-Fe hybrid remains highly bioavailable even when incorporated into polyphenol-rich foods, underscoring its potential as an effective iron fortificant. Fig. 4f and Supporting Fig. S5 show the relative bioavailability of OatNF-SA-Fe, OatNF-NaOH-Fe and FeSO_4_ when administered with water and with açai puree, normalized for each individual participant.

## Discussion and Conclusion

We have introduced in this work a previously unexplored protein-iron nanoparticle hybrid system based on OatNF carrying ultrasmall iron nanoparticles. The synthesis of the OatNF hybrids can be tailored to produce iron nanoparticles in either the ferrous or ferric state; the valence state of the iron nanoparticles is determined by the choice of reducing agent used during synthesis. When sodium ascorbate (SA) is used as the reducing agent, the OatNF carry stabilized ferrous iron which is remarkably well absorbed in humans, with mean fractional absorption of 46% in water, a remarkably 76% higher bioavailability compared to FeSO_4_, the reference compound. When NaOH is used as the reducing agent, the OatNF carry mainly ferric iron, which is still well-absorbed and shows superior sensory performance in reactive food matrices.

OatNF exhibit several key advantages as an iron delivery platform for human nutrition: **i**) the proposed formulation based on oat proteins is suitable for use in all populations, even those with plant-based diets, which typically have both inadequate iron intake and low bioavailability^42,43^; **ii**) OatNF provide efficient iron-binding sites for sub-nm iron particles in both ferrous and ferric form, depending on the reducing agent applied. The binding and alignment of the iron particles is likely due to the intrinsic affinity of proteins to form supramolecular bonds^44,45^. Our results clearly demonstrate that the exceptional bioavailability of iron observed in the OatNF-SA diet is due to the role of protein nanofibrils. Indeed, as evident from the analysis in both solid and dispersion by XPS and colorimetric assays, respectively (Fig.2, Supporting Figs. S4h, S4i), the initial amount of Fe(II) in the oat hybrids and iron sulfate is essentially identical, so that the superior iron bioavailability observed for OatNF-SA hybrids can only be ascribed to the diverse fate of Fe(II) followed during the digestion and adsorption processes of the two formulations; **iii**) OatNF contain abundant reducing amino acids^20^ and a high content of antioxidant peptides^46,47^: these are capable of stabilizing well-absorbed sub-nm ferrous iron nanoparticles in the OatNF-SA-Fe hybrids even during long storage (Fig. 2h); **iv**) the presence of OatNF nanofibrils maintains iron in a colloidally stable state, preventing aggregation and preserving bioavailability; **v**) because of their high solubility in water, their acceptable organoleptic sensory features and pleasant oat flavor and aroma, OatNF-SA-Fe hybrids may be efficient iron fortificants for a wide variety of beverage and food matrices.

In summary, we developed a food-grade oat protein nanofibril-based iron compound for beverage and food fortification that delivers iron with exceptionally high bioavailability. To our knowledge, this is the first study to propose and measure iron absorption from protein-iron nanoparticle hybrid systems in humans. In iron-deficient women, a key target group for iron fortification, the OatNF-SA-Fe hybrid demonstrated exceptional fractional iron absorption. The geometric mean absorption was 46.2% when administered with water and 13.4% when given with a polyphenol-rich food matrix, achieving very high values of bioavailability corresponding to 176% and 166% those of FeSO_4_. Moreover, the OatNF hybrids exhibited sensory performance equal to or better than FeSO_4_ in fortified foods and beverages. These results make OatNF hybrids an innovative and highly effective approach to iron delivery in nutritional applications, which could contribute to reducing the global burden of iron deficiency and anemia.

## Materials and Methods

### Materials

Oat flour (Hafermehl grob, SWISSMILL, Zurich, Switzerland), n-hexane (LiChrosolv®, CAS-No: 110-54-3, Merck KGaA, Darmstadt, Germany), NaOH (BioXtra, ≥98%, pellets, CAS-No: 1310-73-2, Sigma-Aldrich, Buchs, Switzerland), NaCl (≥99.5%, CAS-No: 7647-14-5, Sigma-Aldrich, Buchs, Switzerland). Extracted oat protein, HCl (1.09057, CAS-No: 7647-01-0, Sigma-Aldrich, Reagent European Pharmacopoeia Standard). FeCl_3_, Sodium ascorbate (Sigma-Aldrich 134-03-2, Buchs, Switzerland; Reagent European Pharmacopoeia Standard), NaOH(1.09137, CAS-No: 1310-73-2, Sigma-Aldrich, Reagent European Pharmacopoeia Standard), hydrogen peroxide 30% (1.07298, CAS-No: 7722-84-1, Sigma-Aldrich, Suprapur®) ^54^Fe, ^57^Fe and ^58^Fe enriched elemental iron powders (Chemgas, Boulogne, France). All reagents were of European Pharmacopoeia reagent grade. All steps were carried out with food-grade equipment in the pilot plant/food lab under food-grade conditions. Immediately after the fabrication of this dispersion in Switzerland, it was stored at −20°C for shipping to Thailand.

### Oat protein extraction and fibrillization

The oat protein extraction followed our previous protocol^22^. We also improved the protocol to enhance the yield and productivity showing the same quality nanofibril. Briefly, oat flour was defatted with n-hexane (1:3 w/v) three times for 1 h, centrifuged at 5000 g for 15 min, and air-dried for 24 h. The defatted powder was then dispersed in 1 M NaCl, pH 10 (1:10 w/v) and gently stirred for 2 h at 20 °C before centrifugation (7000 g, 15 min, 20 °C). The protein-rich supernatant was diluted with Milli-Q water (1:6.66 v/v) and left 12 h at 4 ◦C without agitation to precipitate the dissolved oat globulins. After centrifugation (7000 rpm, 15 min, 20 ◦C), the pellet was resuspended in Milli-Q water and dialyzed (MWCO: 6–8 kDa, Spectra/Por RC membrane, Spectrum Laboratories, Rancho Dominguez, CA, USA) against Milli-Q water for 24 h to remove the salt. The dialyzed sample was collected, frozen, and subsequently freeze-dried to obtain the oat globulin (OG) powder.

To fabricate the OG nanofibril, extracted OG was first dissolved in Milli-Q water (pH 2) at a concentration of 2 wt%. After stirring 5 min in ambient condition, the solution pH was then re-adjusted to pH 2. Then, the dissolved protein solution was incubated with a heat treatment (90 °C) by means of an oil/water bath for 18 h while stirring at 350 rpm.

### Production of Oat-SA-Fe and Oat-NaOH-Fe hybrids

For Oat-SA-Fe hybrid, the freshly prepared oat globulin nanofibril solution at the concentration of 2 wt% was first mixed with MQ water and FeCl_3_ solution (0.3 M), and then the freshly dissolved sodium ascorbate (3 M) was added drop by drop, followed by a gentle mixing of the dispersion. The final dispersion reached FeCl_3_ at the concentration of 100 mM and sodium ascorbate at the concentration of 200 mM. For samples for clinical studies, the iron concentration in the dispersion was determined by atomic absorption spectroscopy (AAS). Dispersion with 4mg iron was loaded in each Eppendorf and freeze-dried. The Eppendorf was then filled with N_2_ and then replaced with Argon before shipping for clinical studies.

For Oat-NaOH-Fe hybrid, the freshly prepared oat globulin nanofibril solution at the concentration of 2 wt% was first mixed with MQ water and FeCl_3_ solution (0.3 M) and vortexed for 30s. The dispersion was then carefully adjusted to pH 7 by adding NaOH (7.5M) droplet-by-droplet. The final dispersion reached FeCl_3_ at the concentration of 100 mM. For samples for clinical studies, the iron concentration in the dispersion was determined by atomic absorption spectroscopy (AAS). Dispersion with 4mg iron was loaded in each Eppendorf. The aqueous dispersion was frozen and stored with dry ice before shipping for clinical studies.

The FeCl_3_ solutions (0.3M) were prepared by dissolution of ^57^Fe and ^58^Fe elemental powders in stochiometric amounts of 6 M HCl, followed by oxidation of Fe(II) to Fe(III) with equimolar amounts of 30% hydrogen peroxide.

### Scanning transmission electron microscopy (STEM)

For STEM imaging and analytical analyses, samples were prepared by placing an aliquot of 5 μl on 2nm carbon-coated lacey grids (Quantifoil, D) priorly subjected to glow discharge (Pelco easyGlow, Ted Pella, USA) 2nm carbon-coated lacey grids (Quantifoil, D) for 2 min to ensure the optimal material distribution on the grids. The excess fluid was removed with filter paper and the grids were washed three times with distilled water and air-dried.

The ambient temperature STEM measurements were performed at TFS Talos F200X (Thermo Fisher Scientific, USA), JEOL JEM F200 (JEOL, Japan) instruments both operated at 200kV of acceleration potential and a double C_s_-corrected JEOL JEM-ARM300F GrandARM “Vortex” (JEOL, Japan) instrument operated at 300kV. All three instruments equipped with cold-emission FEG sources were used in a scanning TEM (STEM) mode. For imaging, the high angle annular dark field (HAADF) and circular bright field (BF) detectors were used on all three instruments. The STEM probe of about 0.25 nm in diameter (condenser aperture 70 mm, convergence angle 10.5 mrad) allowed for the time- and position synchronized acquisition of different solid angle ranges of scattered electrons using the coaxially positioned circular bright field and annular dark field detectors. The STEM illumination and acquisition parameters studies were chosen such that the low-angle annular dark field (LAADF STEM) and circular bright field (BF STEM) detectors yielded the diffraction contrast information, whereas the high-angle annular dark field (HAADF STEM) detector provided the prominent atomic number contrast from the same probe position.

### Energy dispersive x-ray spectroscopy (EDS)

The materials elemental content and its distribution in the specimens were assessed by EDS measurements using TFS Talos F200X (Thermo Fisher Scientific, USA) equipped with the SuperX EDS module employing Esprit I package for the evaluation and analyses of spectrum images. The corresponding STEM probe size was set to about 0.25 nm (convergence angle 10.5 mrad, condenser aperture 70 mm) providing the high-resolution imaging and simultaneously yielding sufficient electron probe current for high X-ray count rates. The EDS STEM elemental maps were produced and evaluated using the Esprit I software of the module.

### Electron energy loss spectroscopy (EELS)

The hybridization states of Fe in the compounds were assessed by using EELS modules of both JEOL instruments GrandARM (JEOL, Japan) and JEM F200 (JEOL, Japan). The GIF Quantum ER EELS spectrometer of GrandARM and Gatan Continuum S EEL Spectrometer of JEM F200 allowed the acquisition of EEL spectra in a dual EELS mode. The studies were carried out using 5-7 mA of the emission current and energy dispersion of 0.15eV/channel to include both, O-K and Fe-L23 edges in the spectra.

### CryoTEM

The cryo-TEM samples were prepared in a controlled-environment vitrification system Vitrobot Mark IV (Thermo Fisher Scientific, USA) at 22 °C and 100 % humidity. 3 uL of sample were added on hydrophylised lacey carbon-coated copper grids (EMS, USA), and the excess of sample was blotted with filter paper. Then, the grids were plunge frozen into a mixture of liquid ethane/propane cooled by liquid nitrogen. For automated loading in the cryo-TEM, the vitrified grids were clipped into AutoGrid sample carriers (Thermo Fisher Scientific, USA).

The morphology of the materials in the study was also verified by employing cryogenic transmission electron microscopy (cryo-TEM) studies assess the possibility of electron bean induced material damage. The data acquisition was carried out on a TFS Titan Krios (Thermo Fisher Scientific, USA) operated at 300kV of acceleration voltage and equipped with a Gatan Quantum-LS Energy Filter (GIF) and a Gatan K2 Summit direct electron detector (Ametek Pleasanton, USA). The imaging of the vitrified materials on a cryo-stage constantly kept at 80K was performed in an energy filtered TEM (EFTEM) operation mode using the TFS EPU software and K2 camera in a linear mode.

### X-ray photoelectron spectroscopy (XPS)

The X-ray photoelectron spectroscopy (XPS) measurement was performed in a PHI Quantera system. The powder samples were pressed into In foil before measurement. XPS measurements were performed at a pressure of 10^−9^ − 10^−8^ Torr. The monochromatic Al Kα radiation was generated from an electron beam at a power of 24.8 W with a beam spot diameter of 100 μm. Charge neutralization was performed using a low-energy electron source. Peak fitting of photoelectron features was performed in Casa XPS using Voigt profiles with GL ratios of 40 following Tougaard Background subtraction for the Fe 2p region and Shirley Background subtraction for all other cases. To ensure robustness of the Fe 2p fits especially, literature values reported by A.P. Grosvernor *et al.* were used to constrain the relative peak positions^25^. Relative peak areas between main and satellite peaks were fixed between each analyzed Fe 2p spectra. Furthermore, the full-width half max (FWHM) was fixed to be identical between each main and satellite peaks. The binding energy scale was referenced to the single N 1s feature located at 499.7 eV based on the literature values for amide bondings^48^, thereby using this bonding as the internal charge reference.

### Colorimetric assay

The ferrous and ferric iron content of dispersion was initially investigated by using potassium ferricyanide and potassium ferrocyanide assay. 4% potassium ferricyanide (CAS 13746-66-2, SigmaAldrich) and potassium ferrocyanide (CAS 14459-95-1, Sigma Aldrich) solution were prepared prior to the assay. An aliquot of diluted hybrid dispersion with different concentrations of FeCl_3_ and SA in the plate were mixed with potassium ferricyanide and potassium ferrocyanide solution and waited for 5 min. The plate was transferred to the microplate reader (Infinite M200PRO Tecan) and measured the light absorbance (OD) at 451 nm. The detected absorbances above the detection limit are shown as n.a. in the plot.

The determination of ferrous and ferric iron in our OatNF-SA-Fe dispersion was made by the colorimetric assay using ferrozine and ammonium thiocyanate, respectively. To study ferrous iron, ferrozine (Sigma 160601) in the stock solution (5 mM) was added in the different diluted OatNF-SA-Fe dispersion, reaching the final concentration of 100 µM. The FeSO_4_ and FeCl_3_ solutions at different concentrations and Milli-Q water were used as control. After 10 min, the plate was transferred to the microplate reader (Infinite M200PRO Tecan) to measure the absorbance spectrum and measure the light absorbance (OD) at 562 nm. To study the ferric iron, ammonium thiocyanate (Sigma 221988) stock solution (2 M) was added into the diluted OatNF-SA-Fe dispersion, reaching the final concentration of 300 mM. After 10 min, the plate was transferred to the microplate reader (Infinite M200PRO Tecan) to measure the absorbance spectrum and measure the light absorbance at 480 nm.

### Sensory performance

Color change measurements were performed by adding amounts of Oat-NF-SA-Fe hybrids, Oat-NaOH-Fe hybrids, ferrous sulfate heptahydrate (Sigma Aldrich, Buchs, Switzerland) and ferric pyrophosphate (25% Fe, FCC, Dr. Paul Lohmann GmbH, Emmerthal, Germany) containing 2 mg of Fe to either 250 ml of bottled water (Swiss Alpina Rot, still), UHT apple juice (Pure Apple Juice cloudy still) and UHT chocolate flavored milk (Choco Drink), or 150 g of strawberry yogurt (Naturaplan Organic Strawberry Yogurt), wheat porridge and maize porridges. Wheat and maize porridges were prepared by briefly boiling 100 g of white wheat flour or maize flour in 1 L of water. All products were purchased at the Coop supermarket chain, Zurich, Switzerland. All fortified beverages and foods were prepared in triplicate.

Color change was measured after 2 hours standing at room temperature, compared to the beverages or foods without addition of the iron compounds. Absolute color change (ΔE*_ab_) was determined using a spectral photometer (Chroma Meter CR-410, Konica-Minolta, Tokyo, Japan) in the Hunter Lab color system, calculated as follows:

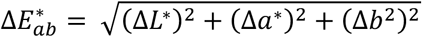

Where ΔL* (lightness), Δa* and Δb* (chromaticity coordinates) correspond to the difference between the sample (with added compound) and the not fortified matrix.

The SPSS statistical programming environment (IBM SPSS Software, Version 28) and Microsoft Office EXCEL 2016 (Microsoft, Redmond, WA) were used for the data analysis. Comparisons were done using two-sided paired t-tests with Bonferroni adjustment for multiple testing. P values <0.05 were considered statistically significant.

### Human study

The study was a single-center, prospective cross-over stable isotope trial, conducted at the Institute of Nutrition of Mahidol University, in Nakhon Pathom, Thailand. The protocol of the study was approved by the Ethical Board of Mahidol University and was registered at clinicaltrials.gov (ID No NCT05826899). All participants provided written informed consent before any study procedure took place.

#### Participants

Women were recruited among students and staff of Mahidol University. The recruitment inclusion criteria were: 1) female; 2) age 18-45 years; 3) serum ferritin (SF) <50 µg/L; 4) body mass index (BMI) 18.5-24.9 kg/m^2^; 5) weight <70 kg; 6) signed informed consent. Exclusion criteria were: 1) anaemia (defined as haemoglobin (Hb) <12 g/dL); 2) presence of thalassemia; 3) inflammation (defined as C-reactive protein CRP > 5 mg/L); 4) cigarette smoking; 5) chronic digestive, renal and/or metabolic disease; 6) chronic medications (except for oral contraceptives); 7) use of vitamin, mineral and pre- and/or probiotic supplements in the previous 2 weeks; 8) blood transfusion, blood donation or significant blood loss in the previous 4 months; 9) history of difficulties with blood sampling; 10) antibiotic treatment in the previous 4 weeks; 11) pregnancy (tested in serum at screening); 12) lactation in the previous 6 weeks; 13) prior participation in a study using stable isotopes or participation in any clinical study in the previous 30 days; 14) unable to comply with study protocol.

#### Study procedures

During screening, about 2 weeks before the start of the study, we collected a venipuncture sample (6 ml) for the determination of Hb, SF, CRP, thalassemia and pregnancy. Body weight and height were recorded, and an interview was conducted to assess inclusion and exclusion criteria. Eligible women were invited to participate to the study.

Each participant received 3 intrinsically-labelled Fe compounds, each administered twice in two study phases: 1) 4 mg Fe as Oat-NF-SA-^57^Fe hybrid 2) 4 mg Fe as Oat-NaOH-^58^Fe hybrid, 3) 4mg Fe as ^54^FeSO4. During the first phase, we administered the compounds on days 1, 3, and 5, diluted in 100 ml reverse osmosis purified water, along with 240 ml purified water as drink. During the second phase of the study (days 19, 21, and 23), the same labelled compounds were re-administered with identical amounts of water, accompanied with 30 g açai puree (The Rainforest Company, Zurich, Switzerland) sweetened with 5 g honey (Doi Kham brand by Royal Doi Kham Food Products Co.,Ltd., Thailand). The order of administration of each compound was randomized for each phase of the study using a Python code. The administrations took place between 07:00 and 09:00 after an overnight fast.

We collected venous blood samples (6 ml) on days 1 (baseline), 19 (phase 1) and 37 (phase 2) to determine Hb, SF, CRP and erythrocyte isotopic composition. Body weight was measured on the same days.

#### Laboratory analysis

Hb, SF and CRP concentrations were measured at the National Healthcare Systems Co., Ltd. (NHealth) in Bangkok, Thailand using an automated hematology analyzer (Sysmex, Kobe, Japan), a chemiluminescent microparticle immunoassay and immunoturbidimetry, respectively. Anemia was defined as Hb <120 g/L. Iron deficiency was defined as serum ferritin <30 μg/L and inflammation was described as CRP >5 mg/L. Whole blood aliquots were sent on dry ice to ETH Zurich, where their iron isotopic composition was measured in duplicate by multi-collector inductively coupled plasma mass spectrometry^49^ (Neptune, Thermo Fisher Scientific, Germany).

#### Sample size

We based our power calculation on data from our previous iron absorption study that administered submicron-sized ferric phosphate in young women in Thailand^19^. In that study, the standard deviation observed between the differences of the logs of the fractional iron absorption (FIA) was 0.223. To resolve a difference of 30% in FIA with a power of 80% and a 5% error rate, we estimated the sample size to be 44 participants. To account for a 20% participant dropout, we recruited 52 participants.

#### Calculations

Fractional iron absorption (FIA) from the iron supplements was calculated based on the shift of the iron isotopic ratios in the collected whole blood samples, using the principles of isotopic dilution and assuming 80% incorporation of the absorbed iron into the erythrocytes^17^. Circulating iron in the body was estimated based on blood volume, derived from body height and weight, and Hb concentration^17^.

#### Statistical analysis

The SPSS statistical programming environment (IBM SPSS Software, Version 28) and Microsoft Office EXCEL 2016 (Microsoft, Redmond, WA) were used for the data analysis. Data were examined for normality by use of the Shapiro–Wilk test. Normally distributed data were reported as mean (SD), and non-normally distributed data were reported as median (IQR). Comparisons were done using two-sided paired t-tests with Bonferroni adjustment for multiple testing. P values <0.05 were considered statistically significant.

## Data Availability

All data produced in the present study are available upon reasonable request to the authors

## Acknowledgements

We thank The Rainforest Company GmbH for financial support and thank Stephan Handschin and Alla Sologubenko from Scientific Center for Optical and Electron Microscopy (ScopeM) at ETH Zurich for the electron microscope imaging and analysis. We thank Michael Sawaya and David Eisenberg from UCLA for the X-ray crystallography analysis of the amyloid structure from a model oat protein hexapeptide fragment. A.W. and S.S. acknowledge funding from the Strategic Focus Area– Advanced Manufacturing (SFA–AM) through the project Advancing manufacturability of hybrid organic–inorganic semiconductors for large area optoelectronics (AMYS).

## Author contributions

R.M. and M.B.Z. conceived the study. J.Z. and R.M. designed and performed the material preparation and their characterization. S.G., C.Z., F.M. and M.Z. designed and performed the clinical study. A.W. and S.S. performed the XPS measurement and analysis, I.K. extracted and purified oat protein. J.Z. C.Z., M.Z. and R.M. wrote the manuscript with input from all authors.

## Ethics declarations

The protocol of the study was approved by the Ethical Board of Mahidol University and was registered at clinicaltrials.gov (ID No NCT05826899). All participants provided written informed consent before any study procedure took place. Mahidol University Central Institutional Review Board (MU-CIRB) is in Full Compliance with International Guidelines for Human Research Protection such as Declaration of Helsinki, The Belmont Report, CIOMS Guidelines and the International Conference on Harmonization in Good Clinical Practice (ICH-GCP. Certificate N° COA No. MU-CIRB 2°023/024.2802.

## Supporting information

**Supporting Figure 1.**
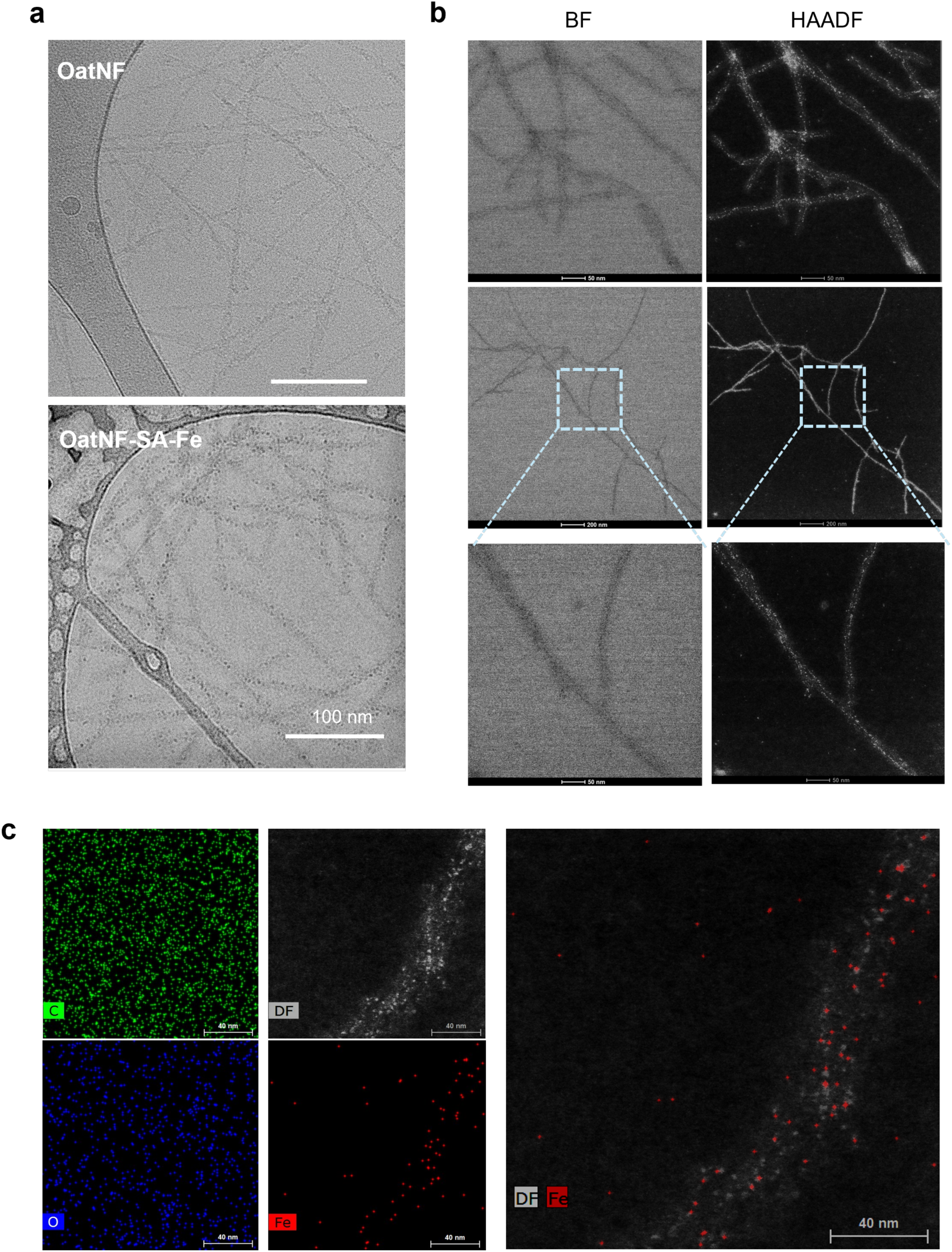
Electron microscopy characterization of synthesized iron fortification OatNF-SA-Fe hybrid. **a**, CryoTEM images of oat nanofibril before and after reducing iron nanoparticles. **b**, STEM images acquired in the high angle annular dark field (HAADF) and circular bright field (BF) detectors. The former provided the prominent atomic number contrast, and the latter yielded the diffraction contrast information. **c**. EDS elemental mapping images of OatNF-SA-Fe hybrids including C, O and Fe, and the corresponding HAADF-STEM image at the same position. The overlay of HAADF-STEM image and EDS elemental map of Fe indicate the accumulation of iron on the surface of oat nanofibril.

**Supporting Figure 2.**
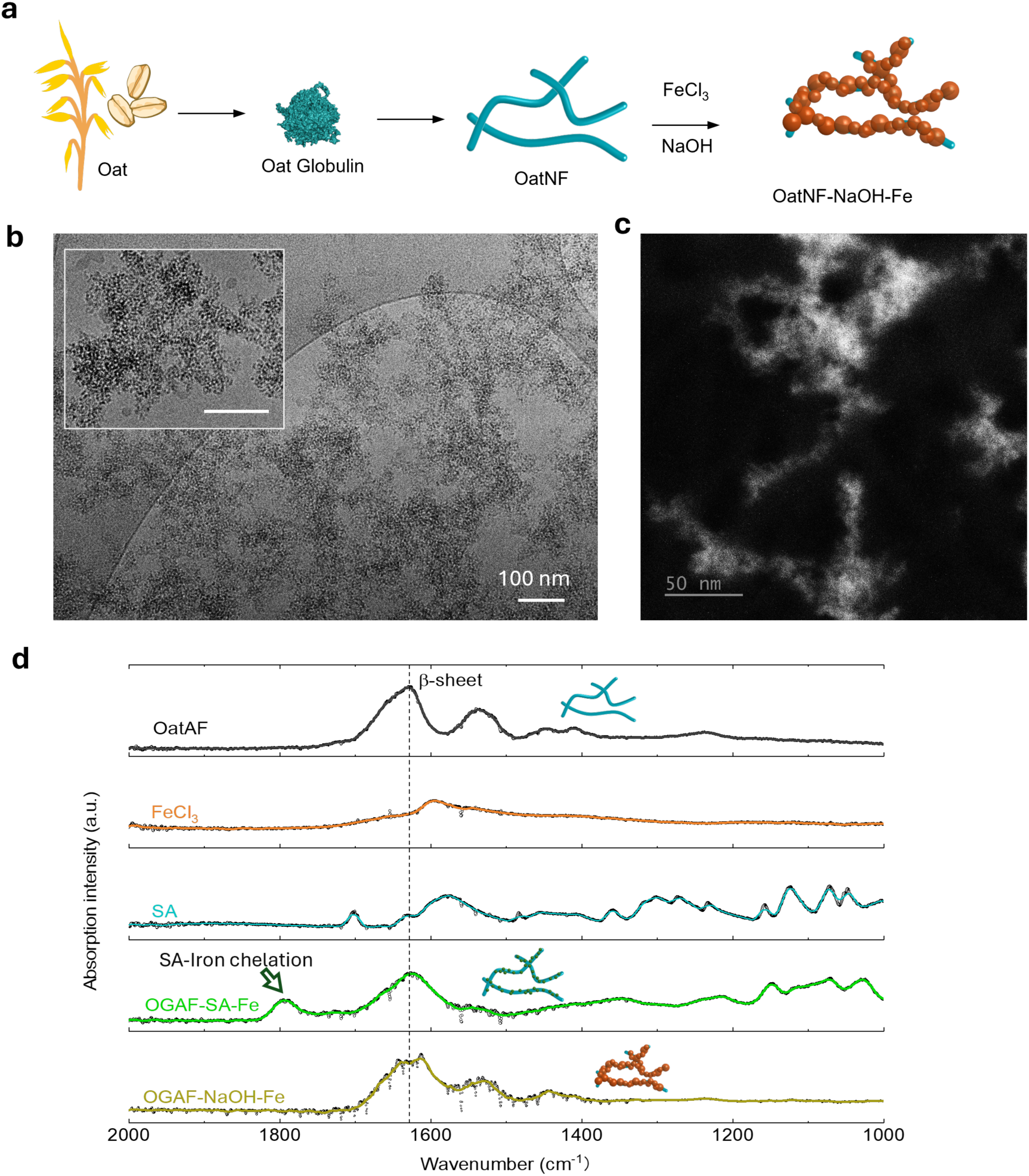
Fabrication of OatNF-NaOH-Fe hybrids and FTIR analysis. **a**, The illustration of fabrication process of OatNF-NaOH-Fe hybrid. **b**, CryoEM images of oat nanofibril of OatNF-NaOH-Fe hybrid **c**. HAADF-STEM image of OatNF-SA-Fe hybrid, where the bright dots indicate the heavier elements that are iron in the hybrid. The rich iron distribution of oat nanofibril is found in this OatNF-NaOH-Fe hybrid, in line with our previous report^1^. (**d**) FTIR analysis of OatNF, FeCl3, SA, OatNF-SA-Fe and OatNF-NaOH-Fe hybrid. The infrared absorption peak of β-sheet (1625 cm^-1^) in the Amide I of oat nanofibril is noted, which are found in both OatNF-SA-Fe and OatNF-NaOH-Fe hybrid. The absorption peak at 1795 cm^-1^ in the OatNF-SA-Fe spectrum denotes the SA-iron chelation, which disappears in the OatNF-NaOH-Fe spectrum. This indicates the effect of SA in preventing iron aggregation and promoting sub-nm iron-binding on the OatNF surface.

**Supporting Figure 3.**
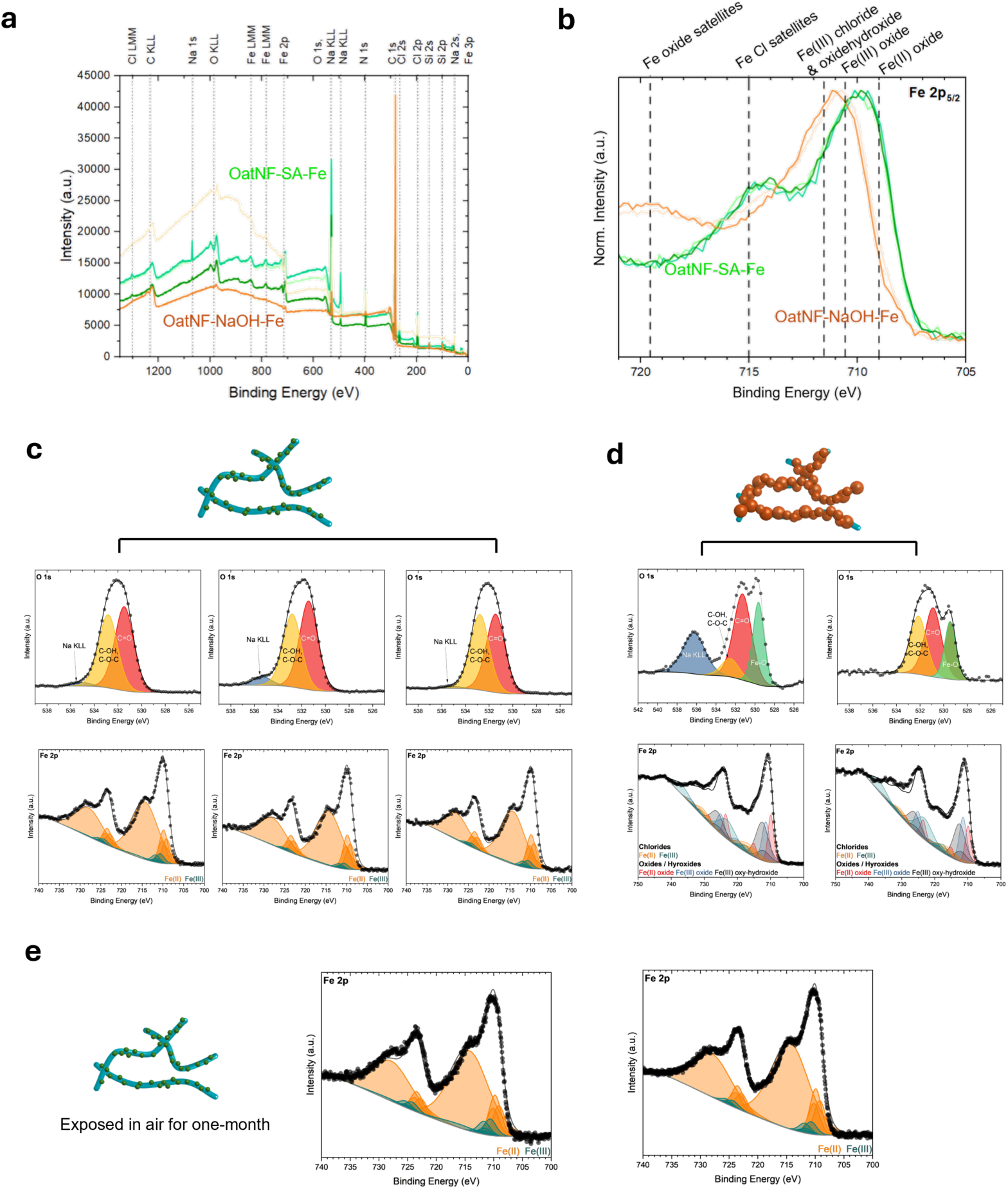
XPS characterization of OatNF-SA-Fe and OatNF-NaOH-Fe hybrids. **a-b**, XPS Survey spectra (**a**) and XPS Fe 2p_5/2_ region (**b**) of OatNF-SA-Fe and OatNF-NaOH-Fe hybrids. The depicted reference lines are based on a previous investigation of the pure compounds by A.P. Grosvernor *et al.*^2^. **c**, Fitted XPS O 1s core level (upper) and fitted XPS Fe 2p core level regions (lower) of OatNF-SA-Fe hybrid, and the. There is no clear Fe-O bond found in the hybrid and the fitting based on literature^2^ showed approximate (92±5)% Fe(II) in the OatNF-SA-Fe hybrid (Fig. 2h). **d**, Fitted XPS O 1s core level (upper) and fitted XPS Fe 2p core level regions (lower) of OatNF-NaOH-Fe hybrids. Clear Fe-O bonds showed in the O 1s spectra indicating the composition of iron oxides and iron oxy-hydroxide. The fitted Fe 2p spectra confirmed these composition and indicated approximately 30-40% Fe(II) species in the OatNF-NaOH-Fe hybrid. **e**, XPS analysis on the OatNF-SA-Fe hybrid after one-month exposure in air. The similar spectra were detected and fitted results revealed (88±5)% and (93±5)% Fe(II) content in the OatNF-SA-Fe hybrid. While these results provide a valuable comparison between samples, they are semi-quantitative in nature.

**Supporting Figure 4.**
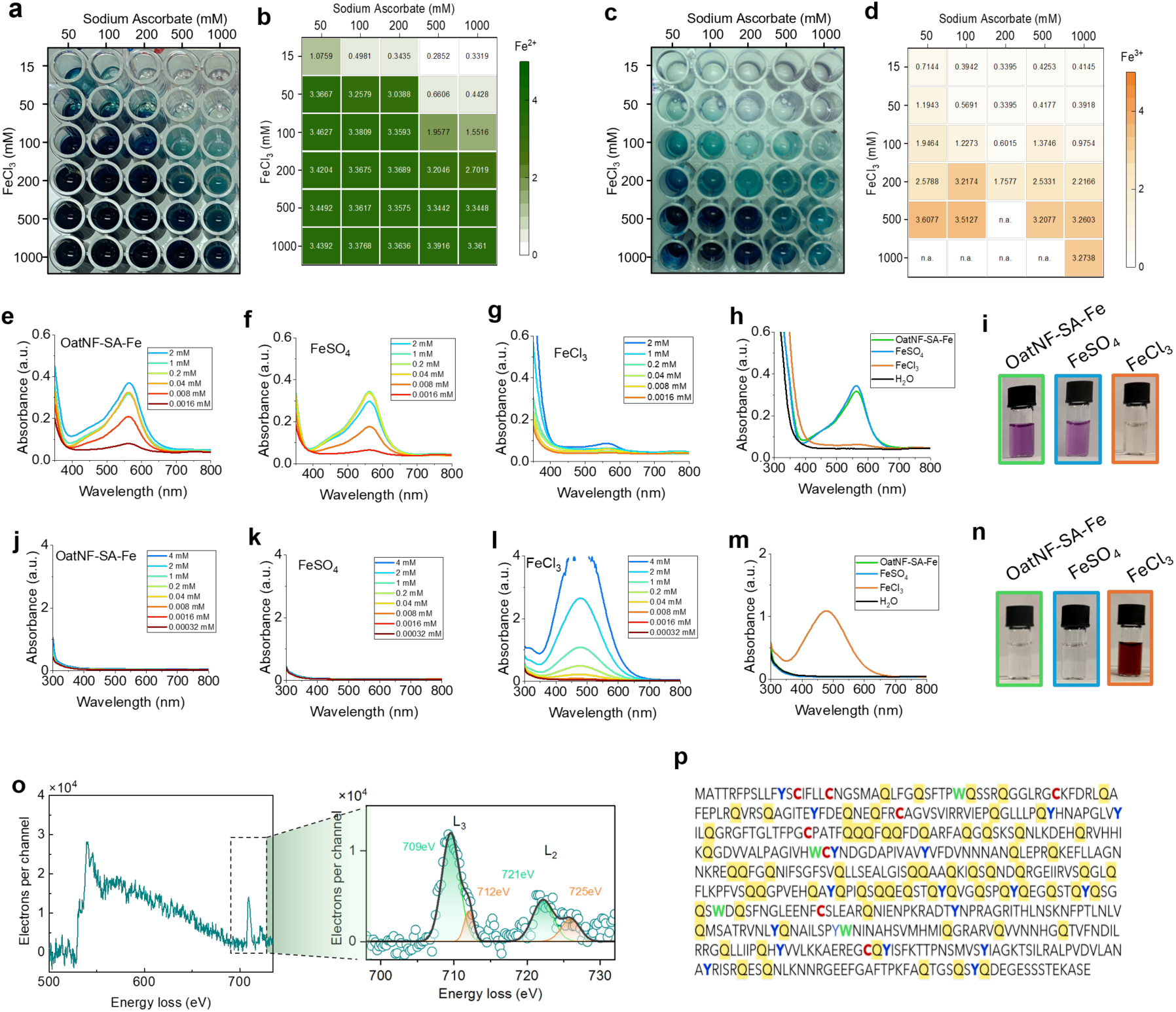
Characterization of OatNF-SA-Fe hybrids. **a-b**, The determination of Fe(II) through potassium ferricyanide colorimetric assay. The photo (**a**) and the absorbance (**b**) using a spectrophotometer of OatNF-SA-Fe dispersion at different SA and FeCl_3_ concentrations were indicated. **c-d**, The determination of Fe(III) through colorimetric assay by potassium ferrocyanide reagent. The photo (**c**) and the absorbance (**d**) using a spectrophotometer of OatNF-SA-Fe dispersion at different SA and FeCl3 concentrations were indicated. (**e-i**) The determination of Fe(II) through colorimetric assay by ferrozine in the OatNF-SA-Fe dispersion (e), FeSO_4_ solution (f) and FeCl_3_ solution (g) at different concentration. Their comparison at Fe concentration of 1mM (h-i) indicates identical, high Fe(II) content in the OatNF-SA-Fe dispersion and FeSO_4_ solution. (**j-n**) The determination of Fe(III) through ammonium thiocyanate colorimetric assay in the OatNF-SA-Fe dispersion (j), FeSO_4_ solution (m) and FeCl_3_ solution (n) at different concentration. Their comparison at Fe concentration of 1 mM (m-n) indicates low Fe(III) content in the OatNF-SA-Fe dispersion and FeSO_4_ solution. (**o**) EELS spectra of our OatNF-SA-Fe hybrids. (**p**) The protein sequence of oat globulin^3^. The amino acids (AAs) with reducing and antioxidant activity^4^ including cysteine, tryptophan and tyrosine are marked, that contribute to the reduction of ferrous iron on the surface of OatNF. The glutamine AA is also highlighted in the protein sequence, account for up to 13% of total AAs, which might promote *in vivo* iron uptake.

**Supporting Figure 5.**
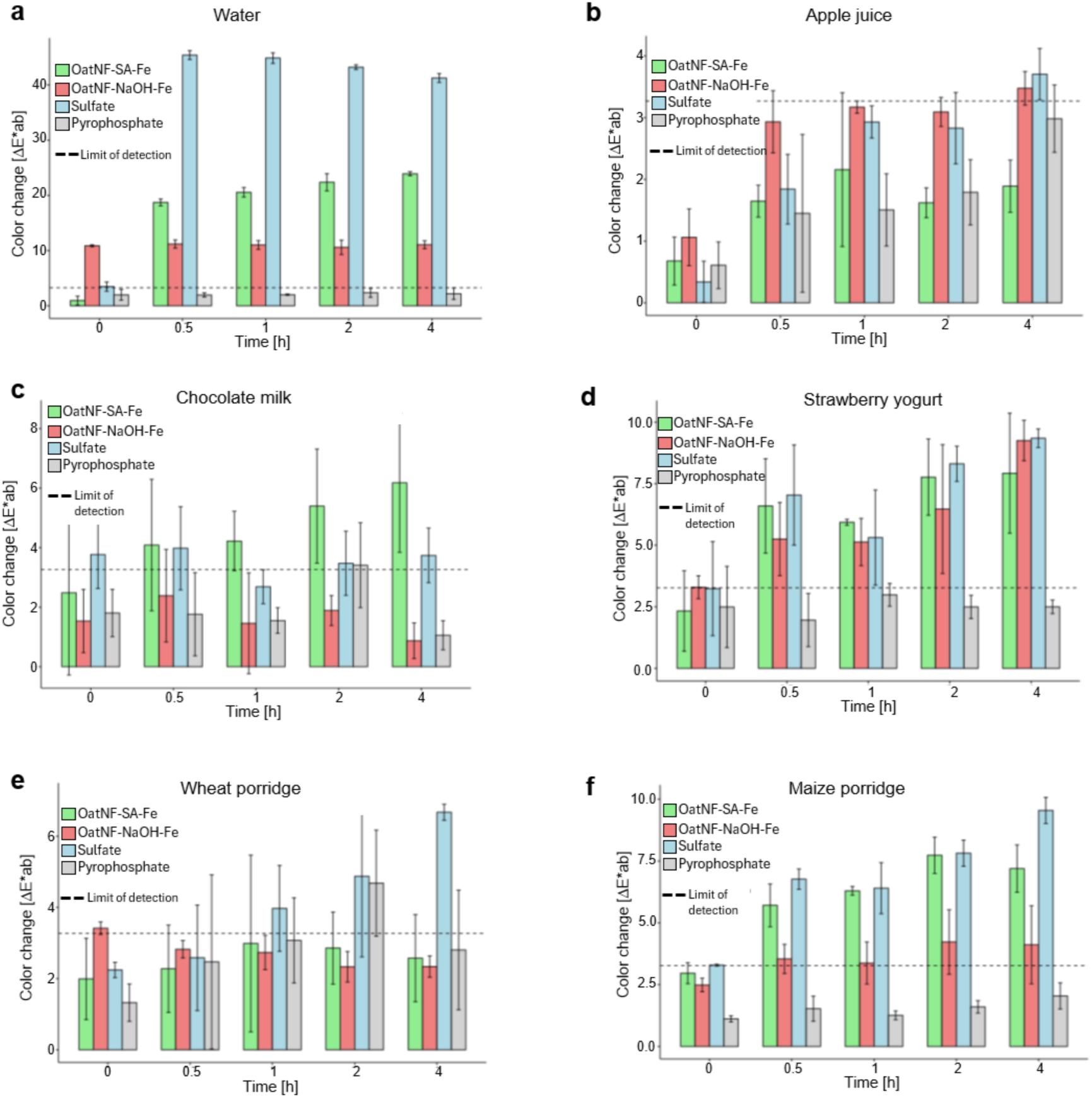
Sensory performance of Oat-SA-Fe, Oat-NaOH-Fe, ferrous sulfate and ferric pyrophosphate in various food matrices. The sensory performance of the OatNF-SA-Fe and Oat-NaOH-Fe hybrids when added to foods and beverages, compared to two commonly used iron fortificants: FeSO_4_ and ferric pyrophosphate, after their addition to bottled drinking water (**a**), apple juice (**b**), chocolate-flavored whole milk (**c**), strawberry yogurt (**d**), and wheat- and maize-based porridges (**e-f**) as a function of time within four hours. N=3 replicates. The data are shown as mean value ± standard deviation. In water, the color change reflects the solubility of the iron compounds and the anti-oxidant effect of SA and nanofibrils, resulting in lower color change for the OatNF-SA-Fe hybrids compared to FeSO_4_. The OatNF-NaOH-Fe fibril exhibits no change over time, reflecting the fact that most of the iron is present as ferric hydroxide/oxide from the start. The food matrices show a similar effect, less pronounced due to the opacity and native color of the matrix, different pH (especially visible for apple juice), buffering effects, and polyphenol contents.

**Supporting Figure 6.**
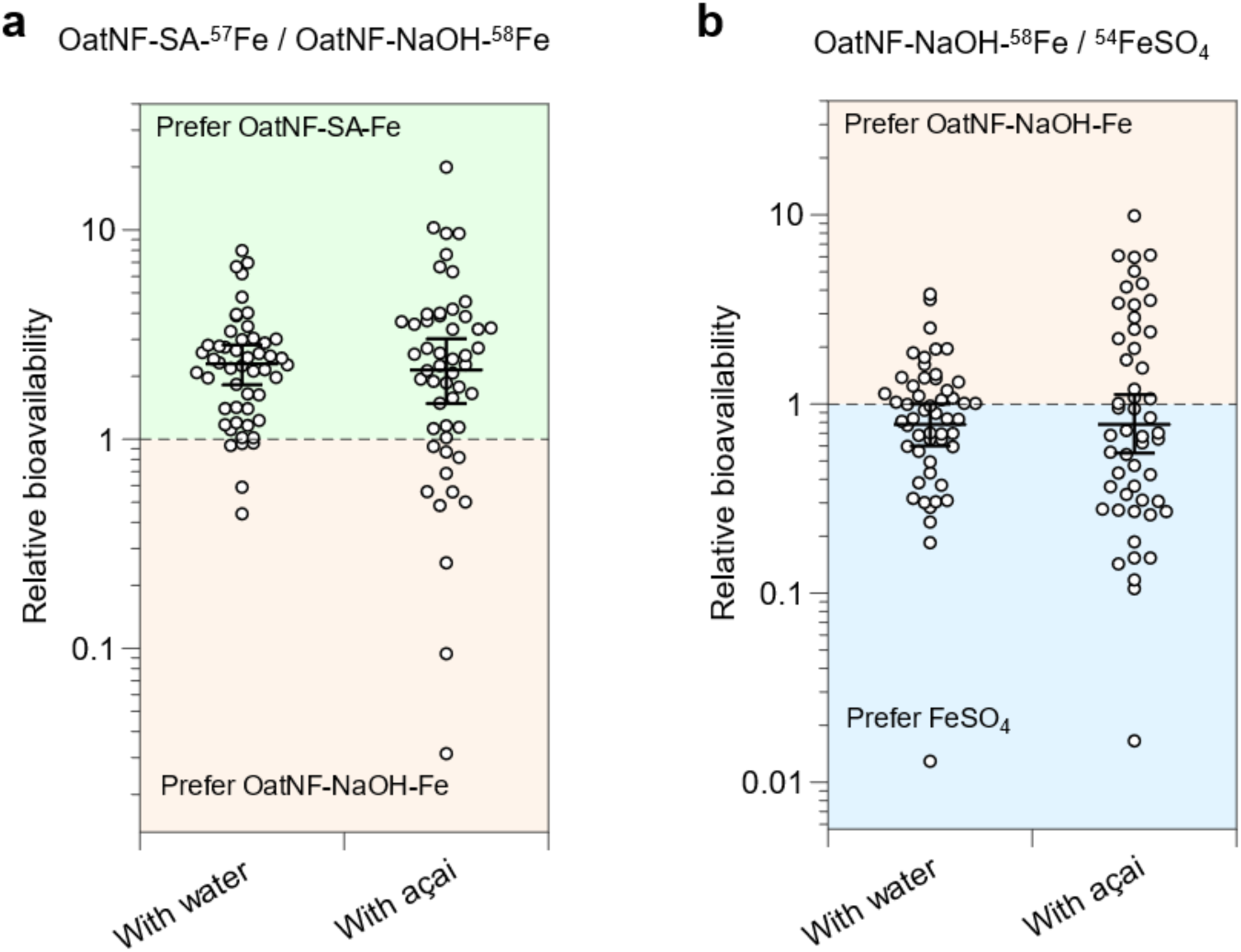
The relative bioavailability among OatNF-SA-^57^Fe hybrids, OatNF-NaOH-^58^Fe hybrids and ^54^FeSO4 according to clinical study. The relative bioavailability between OatNF-SA-^58^Fe hybrids and OatNF-NaOH-^58^Fe (**a**); between OatNF-NaOH-^57^Fe hybrids and ^54^FeSO4 (**b**). The plots show means with whiskers showing SD (n=52).

**Supporting Table 1.** Baseline characteristics of the women participating in the clinical absorption study.

|  |  |
| --- | --- |
| n | 52 |
| Age, y | 32.0 ± 6.8 |
| BMI, kg/m<sup>2</sup> | 21.5 ± 2.0 |
| Haemoglobin, g/dL | 12.80 ± 0.75 |
| Serum ferritin, µg/L | 31.4 (25.1 – 41.3) |
| C-reactive protein, mg/L | 0.73 (0.47 – 1.46) |

Data are mean ± SD or median (IQR).

